# Convergent antibody responses associated with broad neutralization of hepatitis C virus and clearance of infection

**DOI:** 10.1101/2021.09.11.21263416

**Authors:** Nicole E. Skinner, Clinton O. Ogega, Nicole Frumento, Kaitlyn E. Clark, Srinivasan Yegnasubramanian, Andrea L. Cox, James E. Crowe, Stuart C. Ray, Justin R. Bailey

## Abstract

Early development of broadly neutralizing antibodies (bNAbs) targeting the hepatitis C virus (HCV) envelope glycoprotein E2 is associated with spontaneous clearance of infection, so induction of bNAbs is a major goal of HCV vaccine development. However, much remains to be learned at a molecular level about protective E2-reactive antibodies, since HCV infection persists in some individuals despite early development of broadly neutralizing plasma. To examine B cell repertoire features associated with broad neutralization and viral clearance, we performed RNA sequencing of the B cell receptors (BCRs) of HCV E2-reactive B cells of people with cleared or persistent HCV, including subjects with high or low plasma neutralizing breadth in both clearance and persistence groups. We identified many E2-reactive public BCR clonotypes, which are antibody clones with the same V and J-genes and identical CDR3 sequences, shared among subjects grouped by either clearance or neutralization status. The majority (89) of these public clonotypes were shared by two subjects with broad plasma neutralizing activity and cleared infection, but not found in subjects with high plasma neutralizing breadth and persistent infection. We cloned a potent, cross-reactive neutralizing monoclonal antibody (mAb) by pairing the most abundant public heavy and light chains from these two subjects, providing evidence that broadly E2-reactive public clonotypes arise in a subset of individuals with broadly neutralizing plasma and spontaneous clearance of infection. Further characterization of the molecular features and function of these antibodies can inform HCV vaccine development.

## Introduction

Hepatitis C virus (HCV) is a dire global health problem, with an estimated 1.5 million new infections each year and 58 million people living with chronic HCV, a condition which can lead to liver cirrhosis and hepatocellular carcinoma^1^. Although a highly effective cure exists in the form of direct acting antivirals (DAAs), use of DAAs is hampered by limited accessibility, underdiagnosis, and reinfection following cure^2,3^. A prophylactic vaccine is urgently needed to curb the HCV epidemic and help achieve the World Health Organization’s goal of eliminating HCV as a public health problem by 2030^1^.

Approximately 25% of adults spontaneously clear HCV infection, while 75% develop persistent infection^4^. The factors permitting spontaneous clearance of HCV are multifaceted, with important contributions from anti-viral T cells^5-8^ as well as broadly neutralizing antibodies (bNAbs)^9-13^, defined as antibodies that block infection by genetically diverse HCV isolates. In humanized mouse and primate models, bNAb infusion protects against acquisition of HCV^10,11,14^. In humans, early development of high titers of bNAbs is associated with spontaneous clearance^10,11,14^, likely by driving evolution of their target, the HCV envelope glycoprotein E2, to an unfit state^9,15^. However, the precise role of bNAbs in clearance of HCV is incompletely understood. Greater knowledge of molecular features and epitopes of protective bNAbs is still needed, as vaccines that were designed to target HCV envelope proteins have failed thus far to induce high titers of bNAbs^16-18^.

Efforts to understand effective B cell responses in HCV often involve comparisons of people who spontaneously clear HCV and people with persistent infection. However, the dichotomy of clearance and persistence may be insufficient to allow identification of features associated with protective antibodies. Clearance of HCV can occur in the absence of bNAbs, and persistence can occur even when bNAbs are present^19-21^. We previously used a panel of 19 genotype 1 HCV pseudoparticles (HCVpp) to measure neutralizing breadth of plasma from 63 acutely infected individuals who subsequently spontaneously cleared infection or developed persistent infection^10^. Although greater plasma neutralizing breadth was associated with spontaneous clearance of infection, we observed a wide range of neutralizing breadth with both the clearance samples (0-17 HCVpp neutralized) and the persistence samples (0-15 HCVpp neutralized). This observation of relatively high plasma neutralizing breadth in some subjects who failed to clear infection led us to hypothesize that in addition to neutralizing breadth of the plasma polyclonal antibody response, specific molecular features and functions of the monoclonal antibodies (mAbs) making up that polyclonal response may favor clearance or persistence of infection.

We therefore performed an in-depth analysis of the B cell receptors (BCRs) of HCV E2-reactive B cells in people with cleared or persistent HCV, selecting individuals who also demonstrated a range of HCV plasma neutralization capacity to better characterize BCR commonalities between clearance and persistence subjects with both high and low neutralization capacity. We found that grouping of subjects by clearance status or neutralization status revealed differences in V-gene usage and complementarity determining region 3 (CDR3) lengths of BCRs. Most notably, we identified a large number of public clonotypes, which are genetically similar clones of antibodies, shared among subjects grouped by clearance status, neutralization status, or both. The majority of public clonotypes were shared between two subjects with broad plasma neutralizing activity and clearance of infection. Finally, in a proof-of-concept experiment, we produced a potent, cross-reactive neutralizing monoclonal antibody (mAb) by pairing the most abundant public heavy and light chains from these two subjects, suggesting that identification of public BCR clonotypes from individuals selected based on both clearance status and neutralization status can reveal important insights into effective anti-HCV bNAb responses.

## Results

### Selection of subjects

We tested the neutralizing breadth of acute infection plasma samples of 10 subjects from the Baltimore Before and After Acute Study of Hepatitis (BBAASH) cohort with either spontaneous clearance (n=5) or persistence (n=5) of infection who had demonstrated either relatively high or low plasma neutralizing breadth in prior testing^22^, using a well-characterized panel of 19 genotype 1 HCV pseudoparticles (HCVpp)^10^. Plasma samples from clearance subjects were acquired prior to clearance of infection, and samples from persistence subjects were time-matched with the clearance samples for duration of infection. As expected based on our prior study, we observed relatively broad neutralization by plasma samples acquired from some clearance and some persistence subjects. In contrast, plasma from other subjects from each group exhibited very poor neutralizing breadth (Figure 1). Subjects were classified as demonstrating high or low neutralization capacity based on their neutralization score, a previously published metric integrating neutralization potency and breadth^23^, against a panel of 19 genotype 1 HCVpp. When subjects were grouped by neutralization capacity, subjects with high neutralization capacity (neutralization score > 10) had a longer duration of infection (Table 1), as expected based on prior studies showing a correlation between duration of infection and development of neutralizing antibodies^10,11^. Clearance subjects had lower levels of HCV viremia compared to persistence subjects, as expected, since they were sampled at timepoints nearing clearance.

**Table 1.** Subject Characteritics.

|  | Clearance<br>(N=5) | Persistence<br>(N=5) | Significance | High Neut<br>(N=5) | Low Neut<br>(N=5) | Significance |
| --- | --- | --- | --- | --- | --- | --- |
| **HCV Genotype<sup>1</sup>** |  |  |  |  |  |  |
| 1a (%) | 40 | 60 | *ns* | 40 | 60 | *ns* |
| 1b (%) | 20 | 20 | *ns* | 40 | 0 | *ns* |
| 2b (%) | 0 | 20 | *ns* | 0 | 20 | *ns* |
| 3a (%) | 40 | 0 | *ns* | 20 | 20 | *ns* |
| **Duration of**<br>**Infection (Days)<sup>2</sup>** |  |  |  |  |  |  |
| Median | 285 | 318 | *ns* | 344 | 108 | P = 0.02 |
| Range | (82, 379) | (87, 376) |  | (285, 379) | (82, 337) |  |
| **HCV RNA (log<sub>10</sub>**<br>**IU/mL)<sup>2</sup>** |  |  |  |  |  |  |
| Median | 4.4 | 6.5 | P = 0.03 | 6.3 | 5.0 | *ns* |
| Range | (1.4, 6.3) | (5.3, 6.5) |  | (4.4, 6.5) | (1.4, 6.5) |  |

1. Fisher's Exact test was used for HCV genotype comparisons
2. Mann-Whitney test was used for duration of infection and HCV RNA comparisons

**Figure 1.**
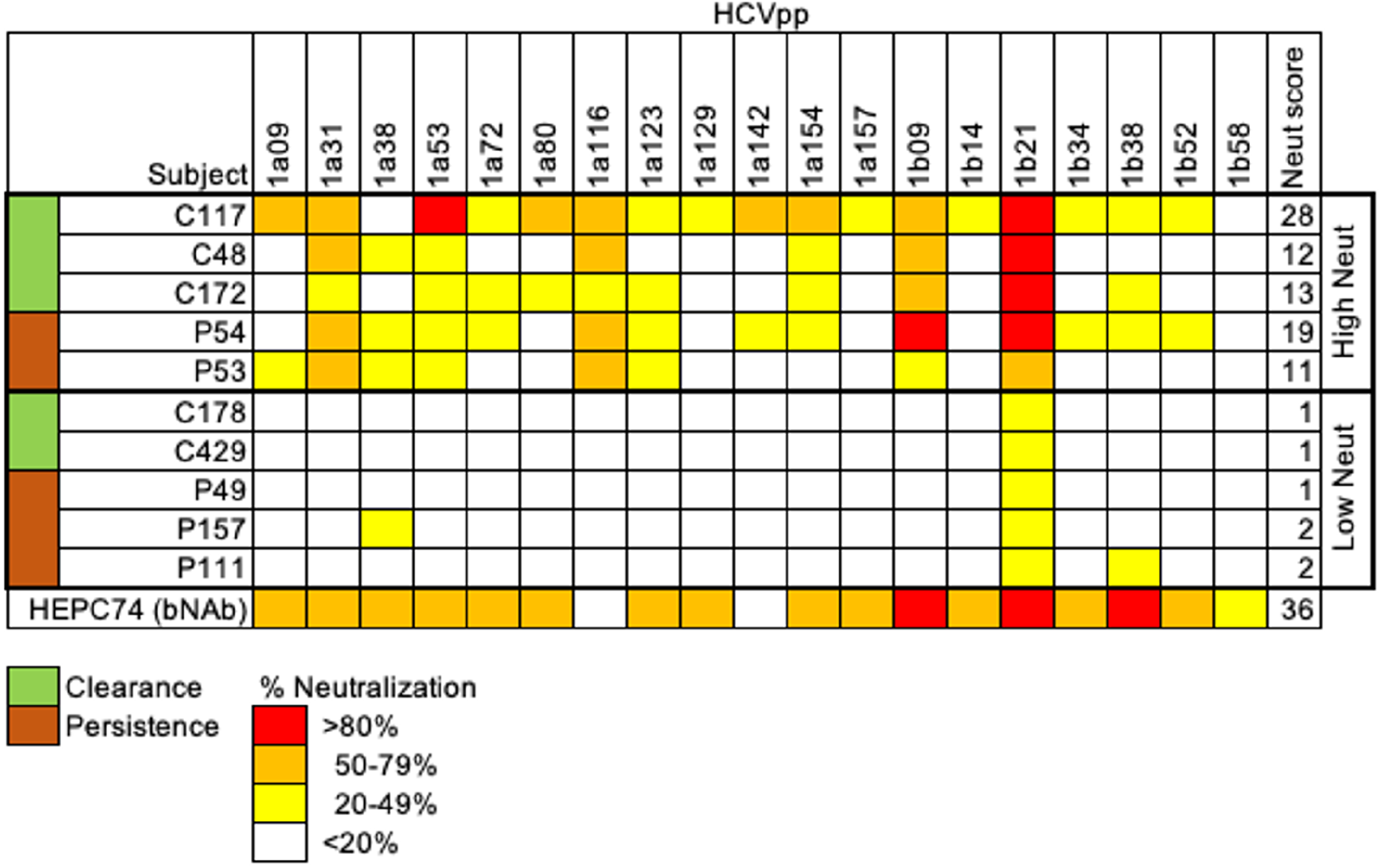
Neutralization score by subject. Percent neutralization achieved by a 1:100 dilution of plasma for each subject was measured using a diverse panel of 19 genotype 1 HCVpp.Values are means of two independent experiments performed in duplicate. As a positive control, the known HCV monoclonal bNAb HEPC74 at 10 µg/mL concentration is shown in the bottom row. Potency of neutralization for each HCVpp is represented by color and breadth by the number of HCVpp neutralized. Neutralization score integrating these two values is shown in the last column, with the score equaling the sum of the potency for each HCVpp (>80% = 3, 50-79% = 2, 20-49% = 1, <20% =0). High neutralization was defined as > 10.

### Sorting of HCV E2-reactive B cells

PBMCs were obtained from subjects at the same timepoints as plasma samples and sorted using flow cytometry to isolate B cells reactive with the HCV E2 envelope glycoprotein. HCV E2-reactivity was determined by binding of class-switched, mature B cells (CD3-, IgD-, IgM-, CD10-, CD19+) to a cocktail of three genotype 1 soluble HCV E2 (sE2) variant proteins (Figure 2a). The three variants (1a157, 1b09, 1b21) were selected because they represented low, medium, or high sensitivity to binding of a panel of previously isolated anti-HCV bNAbs (data not shown). HCV E2 non-reactive B cells were also sorted from each subject as a control. When subjects were grouped based on clearance status, differences were not seen in the frequency of E2-reactive B cells (Figure 2b). However, when subjects were grouped based on neutralization status, subjects with high neutralization scores had a higher frequency of E2-reactive B cells compared to subjects with low neutralization capacity (Figure 2c). Regardless of how subjects were grouped, our assay was sensitive to detect E2-reactive B cells in HCV-infected subjects while not detecting a significant number of false positives in healthy controls (Figure 2).

**Figure 2.**
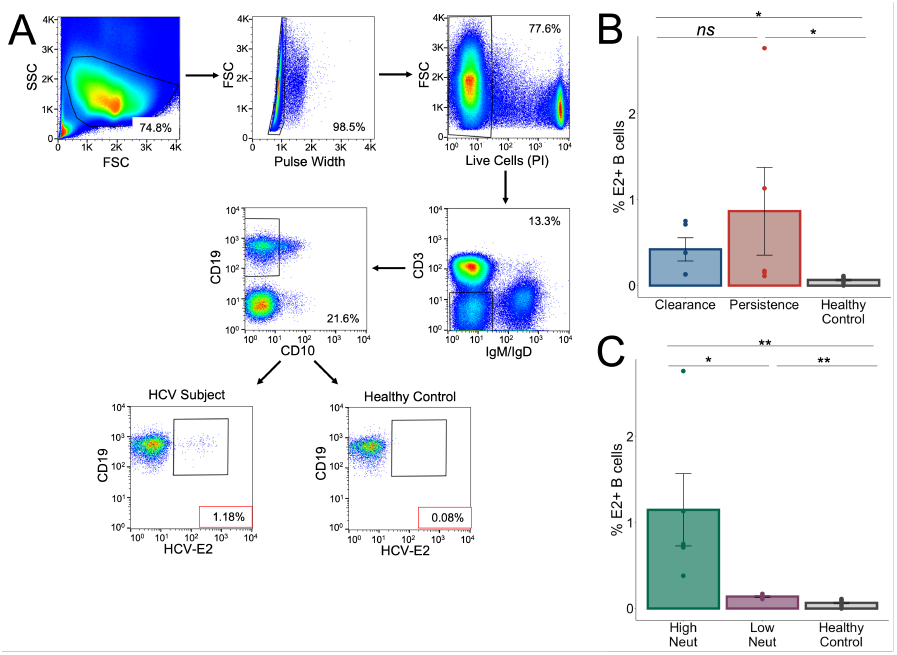
Sorting of E2-reactive B cells. **(A)** Gating strategy is shown for sorting of E2-reactive B cells, showing a representative HCV subject. Final gating of HCV E2^+^ cells is shown in comparison to a healthy control. Frequency of E2^+^ mature, class-switched B cells (% E2^+^ of CD3-, IgM-, IgD-, CD10-, CD19+, live, singlet lymphocytes) is shown for subjects stratified by clearance and persistence **(B)** and by high and low neutralization **(C)**. Lymphocytes were downsampled to 500,000 cells in these analyses so that equivalent numbers of cells in HCV and healthy controls are compared. Statistical comparisons were made using the Kruskal-Wallis test. *, P < 0.05; **, P < 0.01; ns, not significant.

### V-gene usage differs depending on whether subjects are grouped by clearance status or neutralization status

E2-reactive and non-reactive B cells were subjected to bulk RNA sequencing of the variable genes encoding their B cell receptors (BCR-seq). Resultant sequences were grouped into clonotypes using MiXCR^24^, with a clonotype defined as sequences using the same V and J-genes, with identical CDR3 amino acid sequences. For E2-reactive or non-reactive B cells, the number of clonotypes using each V-gene was quantitated for subjects grouped by clearance status and by neutralization status and normalized by the total number of clonotypes identified for each group. A 1.5-fold change in usage with an associated p-value < 0.01 after adjustment for multiple comparisons was deemed to be significant. Comparisons are shown via volcano plot for clearance/E2^+^ compared to persistence/E2^+^ and high neutralization/E2^+^ compared to low neutralization/E2^+^. The pattern of *IGHV* (Figure 3), *IGKV* (Figure 4), or *IGLV* (Figure 5) usage differed depending on whether subjects were grouped by clearance status or neutralization status. For example, *IGHV1-69* had significantly decreased usage in the E2-reactive subset of cells from clearance subjects compared to persistence subjects (p < 1E-20), which was surprising given the large number of mAbs using *IGHV1-69* that have been identified from individuals with clearance of infection^25-27^. However, when stratifying by neutralization capacity, subjects with high neutralization were found to use *IGHV1-69* in their E2-reactive B cells 2.5 times more than subjects with low neutralization (p < 1E-40). The reverse was seen with *IGHV*3-33 which was enriched in clearance relative to persistence E2-reactive B cells but used less frequently in high compared to low neutralization E2-reactive B cells (Figure 3). Although some overlaps were detected in *IGKV* gene use by clearance and high neutralization subjects (e.g., decreased use of *IGKV3D-11, IGKV3D-12, IGKV3D-15*), different genes were enriched in the two groups (Figure 4). Interestingly, there were almost no differences in *IGLV* use regardless of how subjects were grouped (Figure 5).

**Figure 3.**
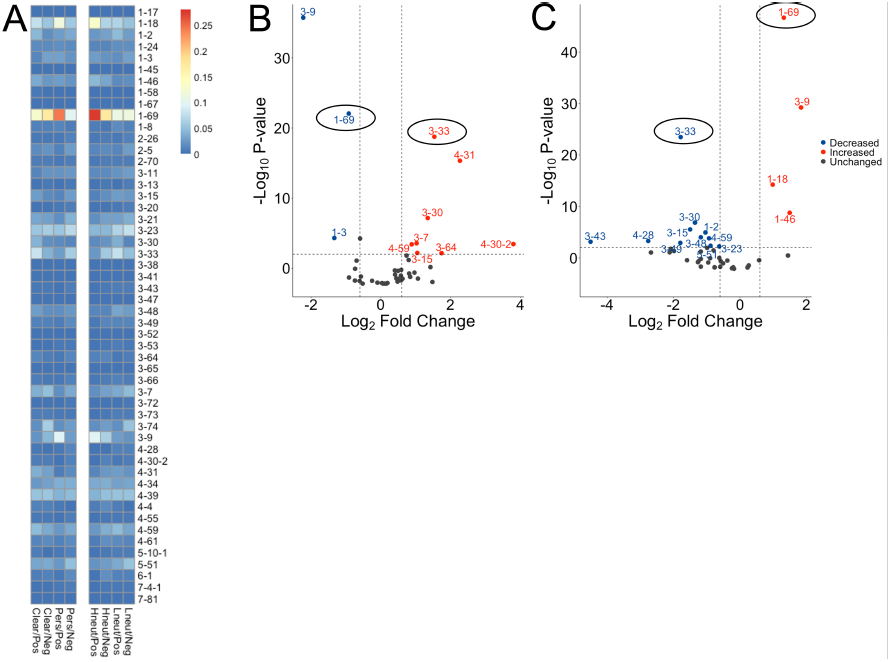
*IGHV* gene usage. **(A)** Heatmap showing *IGHV* gene usage stratified by clearance/persistence (left) or neutralization status (right) of E2-reactive (Pos) and E2 non-reactive (Neg) B cells. Usage of a given *IGHV* gene is expressed as a proportion of total *IGHV* gene usage by the group. Significant differences between E2-reactive clearance and persistence B cells **(B)** and between the E2-reactive high neutralization and low neutralization B cells **(C)** are shown via volcano plot with a significant change defined as a minimum 1.5-fold difference with an associated P value < 0.01 after correction for multiple comparisons. Dashed horizontal lines show the threshold for fold change expressed as Log_2_ Fold Change (0.6 and - 0.6). The dashed horizontal line shows the P value threshold expressed as -Log1_0_ P-value (2). Labeled *IGHV* genes meet significance criteria with those labeled in blue having decreased expression in clearance relative to persistence subjects **(B)** or in subjects with high neutralization relative to low neutralization subjects **(C)**. *IGHV* genes labeled in red have increased expression in clearance or high neutralization subjects. Circled genes are discussed in the text. Statistical comparisons were made using Fisher’s exact test with the Bonferroni correction for multiple comparisons.

**Figure 4.**
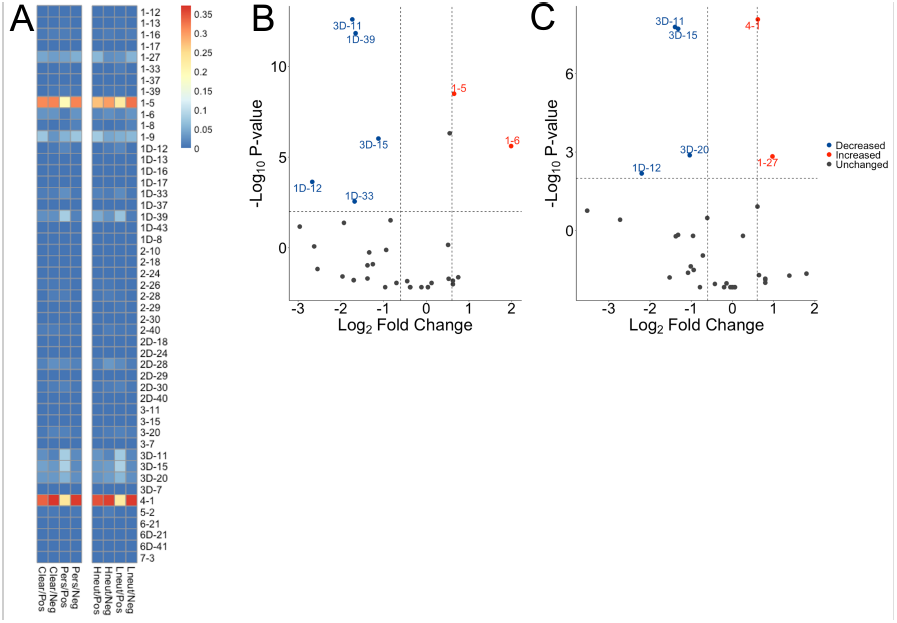
*IGKV* gene usage. **(A)** Heatmap showing *IGKV* gene usage stratified by clearance/persistence (left) or neutralization status (right) of E2-reactive (Pos) and E2 non-reactive (Neg) B cells. Usage of a given *IGKV* gene is expressed as a proportion of total *IGKV* gene usage by the group. Significant differences between E2-reactive clearance and persistence B cells **(B)** and between the E2-reactive high neutralization and low neutralization B cells **(C)** are shown via volcano plot with a significant change defined as a minimum 1.5-fold difference with an associated P value < 0.01 after correction for multiple comparisons. Dashed horizontal lines show the threshold for fold change expressed as Log_2_ Fold Change (0.6 and - 0.6). The dashed horizontal line shows the P value threshold expressed as -Log_10_ P-value (2). Labeled *IGKV* genes meet significance criteria with those labeled in blue having decreased expression in clearance relative to persistence subjects **(B)** or in subjects with high neutralization relative to low neutralization subjects **(C)**. *IGKV* genes labeled in red have increased expression in clearance or high neutralization subjects. Statistical comparisons were made using Fisher’s exact test with the Bonferroni correction for multiple comparisons.

**Figure 5.**
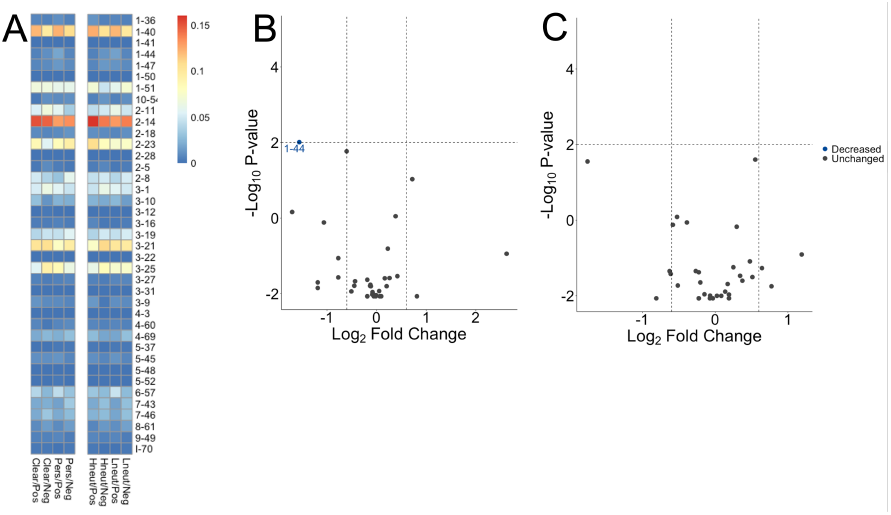
*IGLV* gene usage. **(A)** Heatmap showing *IGLV* gene usage stratified by clearance/persistence (left) or neutralization status (right) of E2-reactive (Pos) and E2 non-reactive (Neg) B cells. Usage of a given *IGLV* gene is expressed as a proportion of total *IGLV* gene usage by the group. Significant differences between E2-reactive clearance and persistence B cells **(B)** and between the E2-reactive high neutralization and low neutralization B cells **(C)** are shown via volcano plot with a significant change defined as a minimum 1.5-fold difference with an associated P value < 0.01 after correction for multiple comparisons. Dashed horizontal lines show the threshold for fold change expressed as Log_2_ Fold Change (0.6 and - 0.6). The dashed horizontal line shows the P value threshold expressed as -Log_10_ P-value (2). Labeled *IGLV* genes meet significance criteria with those labeled in blue having decreased expression in clearance relative to persistence subjects **(B)** or in subjects with high neutralization relative to low neutralization subjects **(C)**. Statistical comparisons were made using Fisher’s exact test with the Bonferroni correction for multiple comparisons.

### CDR3 length differs based on clearance or neutralization status

The amino acid length distribution of CDR3s is often skewed by the response to infection. For instance, bNAbs against HIV often have longer than average heavy chain CDR3s (HCDR3s)^28^. When subjects were grouped by clearance status, HCDR3s were longer in E2-reactive B cells of both the clearance and persistence groups compared to their E2 non-reactive counterparts. However, no difference was detected between the length of HCDR3s in the clearance and persistence E2-reactive B cells (Figure 6a). When subjects were grouped by neutralization status, HCDR3s of E2-reactive B cells of subjects with high neutralization were longer than HCDR3s from subjects with low neutralization (Figure 6d). Minimal differences existed in light chain (LCDR3) length (Figures 6b, c, e, f).

**Figure 6.**
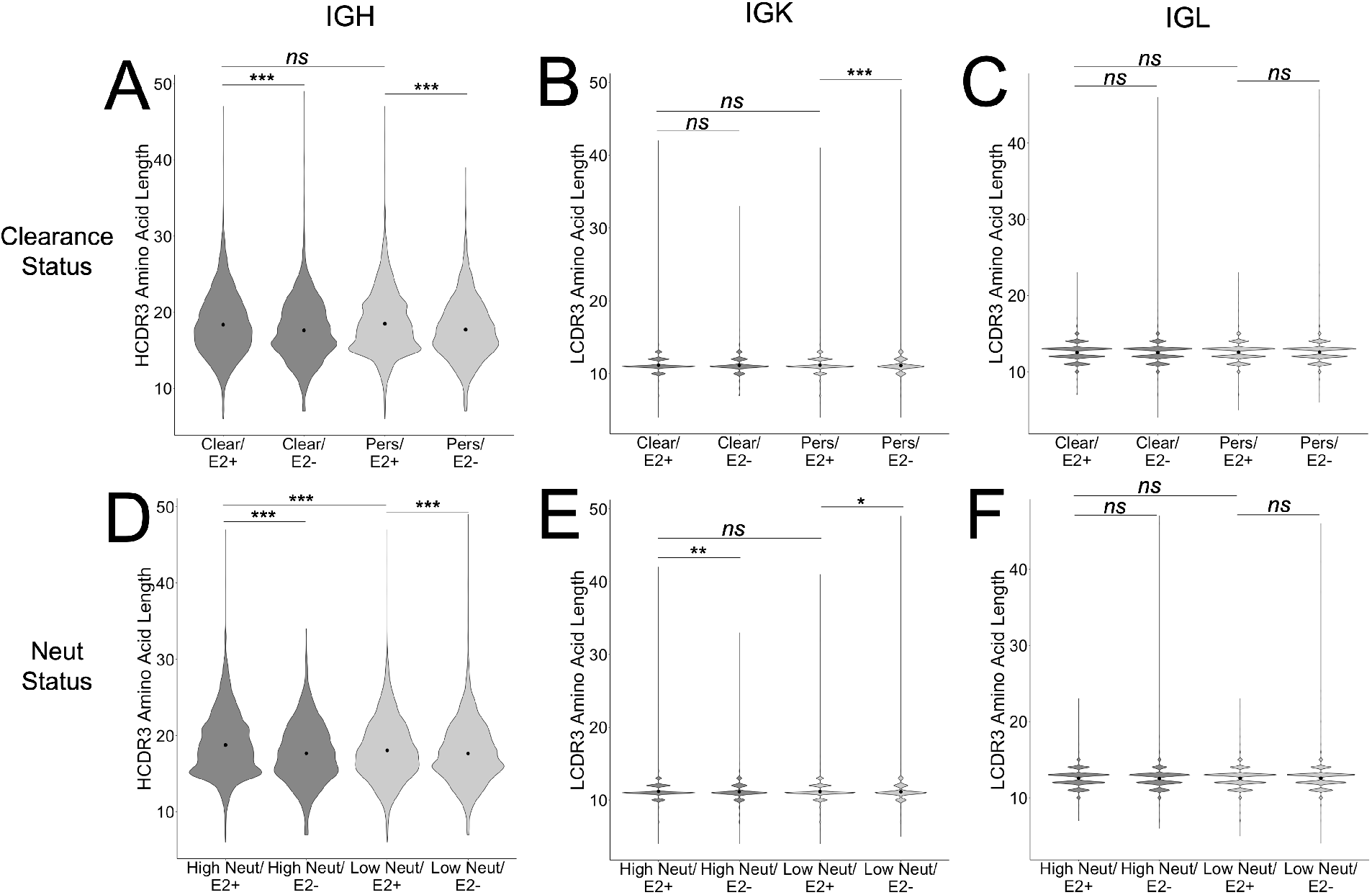
CDR3 length comparisons. Comparison of CDR3 lengths between E2-reactive and non-reactive B cells in clearance and persistence subjects, with analysis of IGH **(A)**, IGK **(B)**, or IGL **(C)** CDR3 sequences. Comparisons are also shown between E2-reactive and non-reactive B cells in high and low neutralization subjects, with analysis of IGH **(D)**, IGK **(E)**, and IGL **(F)** CDR3 sequences. Central dots and vertical lines represent means and standard errors, respectively. Statistical comparisons were made using Kruskal-Wallis test with the Bonferroni correction for multiple comparisons. *, P < 0.05; **, P < 0.01; ***, P < 0.001; ns, not significant.

### HCV E2-reactive public clonotypes were identified from clearance subjects and subjects with high neutralization capacity

Because we were interested in identifying commonalities in BCR sequences favoring the development of bNAbs promoting HCV clearance and broad neutralizing activity, we looked for public clonotypes among clearance subjects and subjects with high neutralization. We defined a clonotype as public if BCRs with identical CDR3 amino acid sequences and the same V and J-gene usage were found in more than one subject of the same group. We further restricted public clonotypes of interest to “group-specific” public clonotypes that were shared only among members of a given group but not present in other groups (e.g., present in multiple subjects in the clearance/E2^+^ group but not present in the clearance/E2^-^, persistence/E2^+^, or persistence E2^-^ groups), increasing the probability of identifying features unique to clearance/HCV neutralization. We found that ∼2.8% of all heavy chain clonotypes among E2-reactive B cells in clearance subjects were group-specific public clonotypes (Figure 7a, left panel). When grouping by neutralization capacity, we found that ∼3.3% of clonotypes of E2 positive B cells of high neutralization subjects were group-specific public clonotypes (Figure 7a, right panel). In contrast, frequencies of groups-specific public clonotypes were lower for E2-reactive B cells of persistence subjects, E2-reactive B cells of low neutralization subjects, or E2 non-reactive B cell subsets (0.4-1.6%).

**Figure 7.**
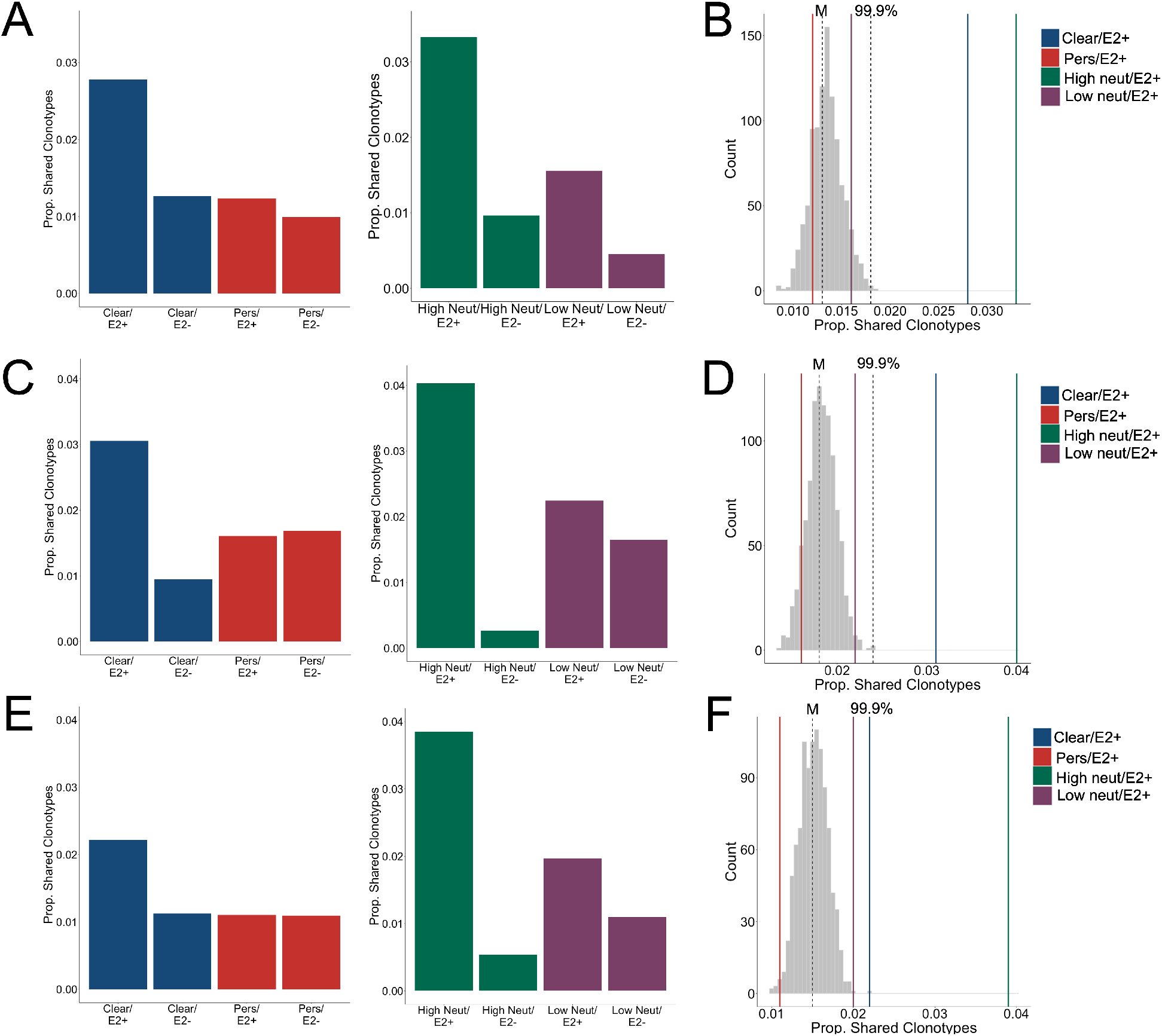
Group-specific public clonotypes. The proportion of group-specific public IGH **(A)**, IGK **(C)**, and IGL **(E)** clonotypes for subjects stratified by clearance status (left) or neutralization status (right) is shown. A histogram shows the frequency of group-specific public clonotype proportions resulting from 1000 trials of random permutation of the IGH **(B)**, IGK **(D)**, and IGL **(F)** clonotype data. Medians are marked by dashed vertical lines labeled with an “M” and the 99.9^th^ percentile is marked with a dashed vertical line labeled with “99.9%.” The proportion of group-specific public clonotypes used by clearance/E2^+^, persistence/E2^+^, high neutralization/E2^+^, and low neutralization/E2^+^ groups are marked with solid, color-coded vertical lines. In **(F)**, The 99.9^th^ percentile overlaps with the line denoting the proportion of clearance/E2^+^ public clonotypes. Public clonotypes are expressed as the proportion of clonotypes sharing an identical CDR3 amino acid sequence and using the same V and J-genes out of the total number of clonotypes in the group.

To compare these frequencies with the amount of clonotype sharing that would be expected due to chance, we performed random permutation of data, wherein clonotypes were randomly reassigned to subjects irrespective of group, and the frequency of group-specific public clonotypes for a group of 5 subjects was calculated. This process was performed 1000 times and a probability distribution of group-specific public clonotype frequencies was generated (Figure 7b). The median of this distribution for heavy chain clonotypes was 1.3% with the 99.9^th^ percentile falling at 1.8%. The frequencies of group-specific public clonotypes for E2-reactive persistence, E2-reactive low neutralization, and all E2 non-reactive groups fell within this distribution. However, for E2-reactive B cells in clearance subjects or E2-reactive B cells in high neutralization subjects, frequencies of group-specific public clonotypes fell well above the 99.9^th^ percentile, making it exceedingly unlikely that this degree of clonotype sharing would occur by chance. We performed the same analysis for IGK (Figure 7c-d) and IGL (Figure 7e-f) with similar results, noting only that with IGL, the frequency of group-specific public clonotypes for the E2-reactive clearance group fell at the 99.9^th^ percentile. We analyzed V-gene usage (Supplemental Figure 1) and CDR3 length (Supplemental Figure 2) among the public clonotypes, but did not detect any notable differences between clearance/persistence or high/low neutralization subjects.

### Unbiased clustering of subjects based on clonotype sharing

Because both clearance status and neutralization status could drive sharing of clonotypes, we investigated how sharing occurred between subjects in a non-biased way. We quantitated the number of shared clonotypes between each pair of subjects regardless of group and performed average-link clustering to reveal relationships among subjects. When analyzing sharing of clonotypes between E2-reactive B cells (Figure 8a), we found that subjects tended to cluster by their neutralization status rather than by their clearance status, with only one outlier (subject P157). However, the highest degree of sharing by far occurred between a single pair of subjects (C48 and C117) who were both clearance and high neutralization subjects. These two subjects also had the highest proportion of shared IGK and IGL clonotypes (Figure 8a, middle and right panels, respectively). Infecting HCV genotype did not explain clonotype sharing of E2-reactive B cells, as subjects with the most sharing did not share an infecting genotype. Most notably, C48 or C117 were infected with genotype 3a or 1a virus, respectively. In contrast, the E2 non-reactive B cell clonotype clustering occurred without segregation by either clearance or neutralization status and appeared to be more strongly related to infecting HCV genotype (Figure 8b).

**Figure 8.**
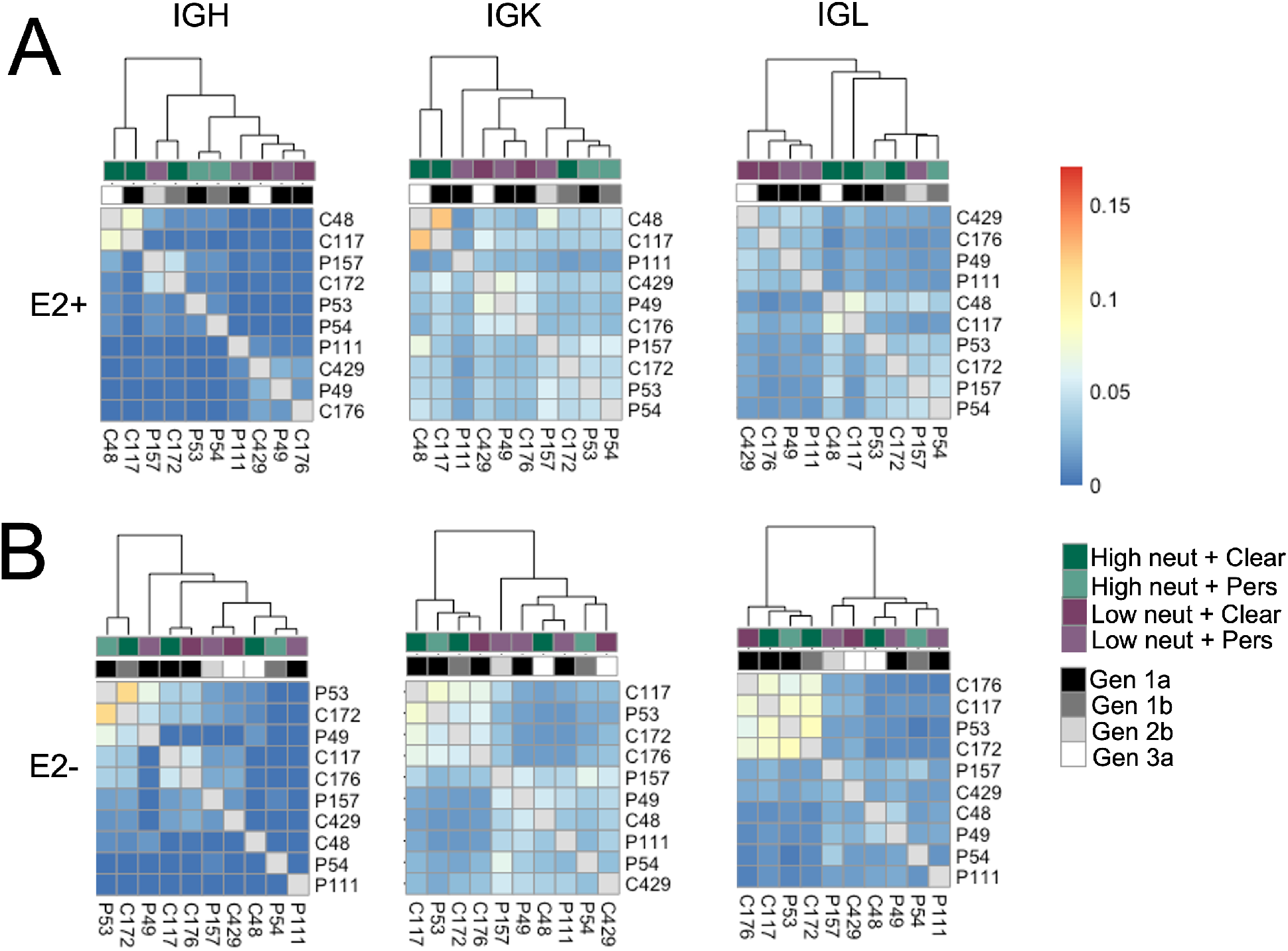
Unbiased clustering of subjects based on clonotype sharing. Heatmaps showing sharing of IGH (left), IGK (middle), and IGL (right) clonotypes from E2-reactive **(A)** and E2 non-reactive **(B)** B cells between all subjects. Each column and row are labeled with a subject. Each column also has a colored box to indicate that subject’s clearance and neutralization status as well as a grayscale box to indicate that subject’s infecting HCV genotype. The ordering of subjects is determined by average-link clustering with the resultant dendrogram shown at the top of each heatmap.

### Antibody production and neutralization analysis

The large number of public clonotypes shared by C48 and C117 (89 IGH, 110 IGK, and 70 IGL clonotypes) were of interest since they derived from subjects at the overlap of clearance and high neutralization capacity. In addition, prior work we have done on the repertoire of subject C117 revealed that this subject produced broadly neutralizing antibodies associated with evolution of HCV to an unfit state, leading to clearance^9^. Consequently, we hypothesized that these shared sequences encoded anti-HCV bNAbs. Because we performed bulk RNA seq analysis of heavy and light chain variable genes separately, we could not determine the authentic pairing of BCR heavy and light chains. However, as a proof of principle experiment, we produced a mAb by cloning and expressing the most abundantly expressed public heavy chain and most abundantly expressed public light chain (IGK) sequences that were shared by these subjects (Figure 9a). This mAb bound to three different variant E2 proteins with an affinity comparable to a control bNAb, HEPC74 (Figure 9b). Although this antibody was not broadly neutralizing, it exhibited >50% neutralization for 5 of 17 HCVpp in a panel with four tiers of increasing neutralization resistance, including both genotype 1 and genotype 5 isolates (Figure 9c).

**Figure 9.**
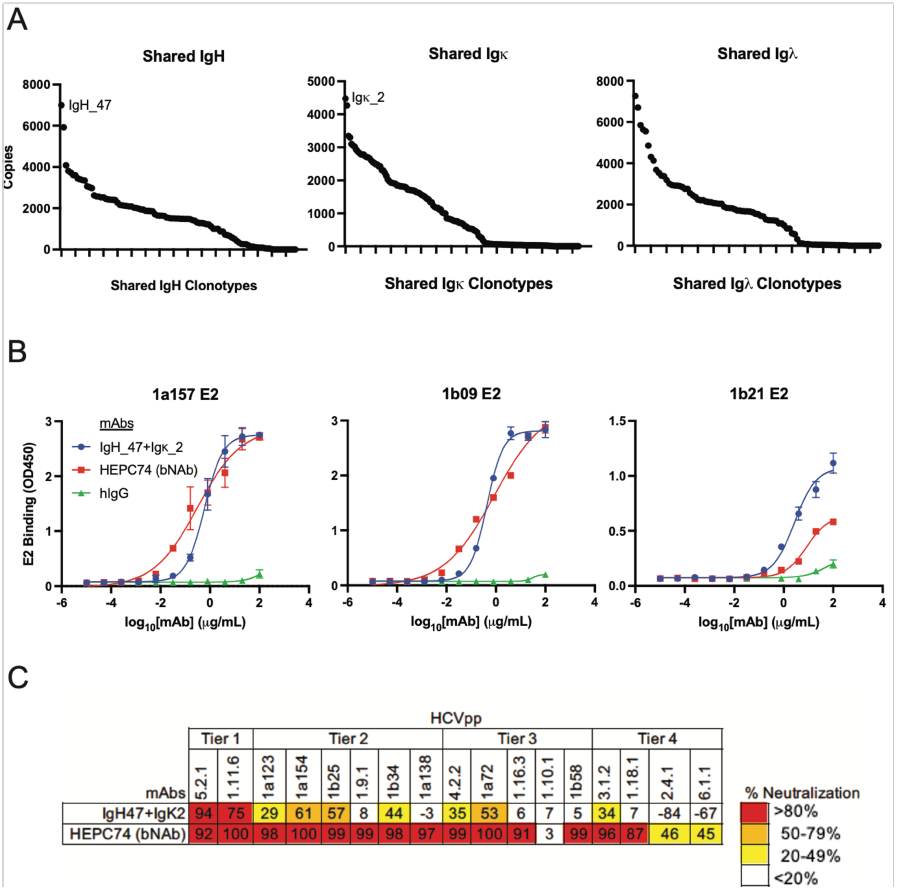
Binding and neutralization assays for public clonotype mAb. **(A)** Abundance of group-specific public clonotype CDR3 sequence copies is shown for IGH (left), IGK (middle), and IGL (right). The most abundant IGH public clonotype (designated IGH_47) and the most abundant light chain public clonotype (designated IGK_2) were cloned into IgG1 and IgK expression vectors and co-transfected to generate a mAb. **(B)** E2 binding ELISA of the public clonotype mAb (IGH_47 + IGK_2, blue) using 1a157 (left), 1b09 (middle), and 1b21 (right) E2 variant proteins. The HEPC74 bNAb (red) was used as a positive control and non-reactive human IgG (green) was used as a negative control. (C) Neutralization breadth of the public clonotype mAb (IGH_47 + IGK_2) measured at 100 µg/mL concentration using a panel of 17 genotype 1-6 HCVpp selected to span 4 tiers of increasing neutralization resistance. Values are the average of duplicate wells. As a positive control, the known HCV bNAb HEPC74 tested at 100 µg/mL concentration is shown in the bottom row.

## Discussion

We sorted E2-reactive B cells from subjects with acute HCV infection prior to clearance and from subjects with persistent infection time-matched with the clearance subjects for duration of infection. These subjects were also selected to represent individuals with high or low plasma neutralization activity (Figure 1). Subjects with high neutralization capacity had higher frequencies of E2-reactive B cells compared to low neutralization subjects (Figure 2). Similarly, differences in HCDR3 length emerged between BCRs of E2-reactive B cells from high or low neutralization subjects that weren’t present between E2-reactive cells of clearance or persistence subjects. Stratifying subjects by neutralization capacity also yielded different patterns of V-gene usage (Figures 3-5) compared to stratifying by clearance status. For example, *IGHV1-69*, a gene used by many HCV bNAbs^25-27^, was less frequently used in E2-reactive B cells of clearance subjects relative to persistence subjects, but this V-gene was enriched in high neutralization relative to low neutralization subjects. Therefore, stratifying by neutralization status or clearance status alone would have provided an incomplete picture of potentially important BCR molecular features.

This conclusion was underscored by our analysis of public clonotypes. We identified group-specific public clonotypes shared among E2-reactive B cells of clearance subjects or E2-reactive B cells of high neutralization subjects (Figure 7). An unbiased clustering analysis showed that overall sharing among subjects was driven more by neutralization status than clearance status, but also showed that the most sharing by far occurred between subjects C48 and C117 who were both clearance and neutralization subjects (Figure 8). In total, a surprisingly high percentage of the clonotypes in the samples from the individuals C48 and C117 (∼10% or 16% of IGH clonotypes, respectively) were public clonotypes shared with one another. To our knowledge, this is the first identification in HCV of public B cell clonotypes meeting this high degree of similarity. Unfortunately, although we attempted to identify V-gene usage patterns (Supplemental Figure 1), CDR3 length similarities (Supplemental Figure 2), or sequence motifs (data not shown) in the group-specific public clonotypes of clearance or persistence subjects, or subjects with high or low neutralization capacity, we were unable to do so. This may be because such motifs are complex or non-linear, or because although shared, the public clonotypes are diverse.

Since these experiments were done using bulk RNA seq analysis, we did not know the authentic pairing of heavy and light chain sequences. However, we synthesized an antibody using the most abundant shared IGH and light (IGK) chains from C48 and C117, hypothesizing that since both subjects cleared HCV and produced neutralizing antibodies, their shared BCR sequences had a high probability of yielding neutralizing antibodies. Despite the significant risk of mispairing of the heavy and light chain from such an approach, the antibody produced was functional, potently binding to diverse E2 proteins (Figure 9b) and even potently neutralizing numerous HCVpp (Figure 9c). This finding provides proof of concept of the potential benefits of BCR analysis focused on E2-reactive B cells from subjects stratified by infection outcome and plasma neutralizing activity. Using single-cell RNA-seq analysis to obtain paired heavy and light chain data is a potential next step to enable in-depth functional analysis of the public clonotypes from individuals with broad neutralizing activity and clearance of HCV infection.

## Materials and Methods

### Study subjects

PBMC and plasma samples from HCV-infected individuals were obtained from the Baltimore Before and After Acute Study of Hepatitis (BBAASH) cohort^29-31^. This cohort was established at Johns Hopkins Medicine more than two decades ago and is a prospective cohort of injection drug users in Baltimore who are followed from before infection with HCV, through spontaneous clearance/persistence of HCV over as many as 15 years. The study was approved by the Institutional Review Board of Johns Hopkins Hospital, and informed written consent was obtained from all study participants.

### HCV viral load and clearance/persistence determinations

HCV viral loads (IU/mL) were quantified after RNA extraction with the use of commercial real-time reagents (Abbot HCV Real-time Assay) migrated onto a research-based real-time PCR platform (Roche 480 LightCycler). HCV seropositivity was determined using the Ortho HCV version 3.0 ELISA Test System (Ortho Clinical Diagnostics). Subjects were assigned to either clearance or persistence groups based on infection outcome. Clearance was defined as undetectable HCV RNA for a period of at least 60 days with no recurrence of viremia in individuals with detectable anti-HCV antibodies. Persistence was defined as detectable HCV RNA viremia for more than 1 year. Day 0 of infection was estimated as the midpoint date of the last negative and first positive HCV RNA test. Duration of infection was calculated as that timepoint’s date minus the date of day 0.

### HCVpp neutralization assay and high/low neutralization capacity determinations

HCVpp were produced by lipofectamine-mediated transfection of HCV E1/E2, pNL4-3.Luc.R-E, and pAdVantage (Promega) plasmids into HEK293T cells as previously described^32,33^. Neutralization assays were performed as described previously^34^. mAbs at 10 or 100 μg/mL, or heat-inactivated plasma samples at 1:100 dilution were incubated with HCVpp for 1 hour at 37°C prior to addition to Hep3B cells in duplicate. Medium was changed after 5 hours, and cells were incubated for 72 hours before measuring luciferase activity in cell lysates in relative light units (RLUs). Only HCVpp preparations producing RLU at least 10-fold above values of mock pseudoparticles were used for neutralization experiments, and HCVpp input was normalized to 1 × 10^6^ to 6 × 10^6^ RLUs. Nonspecific human IgG (Sigma-Aldrich) was used as a negative control. To quantitate neutralization capacity, integrating both breadth and potency, we used a neutralization score^23^ whereby plasma neutralization potency was scored for each HCVpp (>80% neutralization received a score of 3, 50-80% received a score of 2, 20-49% received a score of 1, and <20% received a score of 0) and then summed over all HCVpp, using a previously characterized panel of 19 HCVpp representing diverse HCV E1E2^10^. We defined high or low neutralization subjects as subjects with neutralization scores > 10 or < 10, respectively.

### Generation of soluble E2 (sE2) for B cell sorting

Genes encoding E2 ectodomains of three genotype 1 HCV strains (1a157, 1b09, 1b21) were cloned from a previously described library of E1E2 clones^35^ into a mammalian expression vector (phCMV3-Ig Kappa-HIS, a gift of Leopold Kong, The Scripps Research Institute, La Jolla, California, USA). The vector allows expression of soluble E2 protein with a C-terminal His tag as well as an N-terminal murine Ig Kappa leader signal for efficient protein secretion. sE2 expression and purification was performed as previously described^9^.

### Cell staining and flow cytometric cell sorting

PBMCs were isolated from blood using a Ficoll separation gradient. 30-50×10^6^ PBMCs were incubated with anti-CD81 antibody (BD Cat #555675) at 5ug/ml and Fc blocker (BD Cat #564220) diluted in FACS Buffer (1x PBS with 1% BSA) for 30 minutes at 4ºc. Cells were then washed twice with FACS buffer. sE2 cocktail (1a157, 1b09, and 1b21) was added to the cells at 5ug/ml and incubated at 4ºc for 30 minutes. Cells were washed three times with FACS buffer. Fluorophore conjugated antibody cocktail containing CD10-PE (BD Cat #555375), CD19-BV421 (BD Cat #562440), CD3-APC H7 (BD Cat#560176, IgM-BB515 (BD Cat #564622), IgD-BB515 (BD CAT #565243), and Anti His-AF647 (ThermoFisher Cat #MA1-21315-A647) was added to the cells. They were incubated at 4ºc for 30 minutes then washed three times. The viability dye propidium iodide (PI) was added immediately prior to sorting. Cells were sorted using a MoFlo Legacy cell sorter (Beckman Coulter). Mature, class switched Bells were gated as follows: lymphocytes (FSC by SSC), singlets (FSC by pulse width), live cells (PI), IGM-, IgD-, CD3-, CD10-, CD19+. E2^+^ and E2^-^ B cells were sorted directly into cell lysis buffer (Qiagen). PBMCs from at least one healthy control subject were stained with each cell sorting experiment and used for setting of the E2^+^ gate. Nine to ten thousand E2-non-reactive (E2^-^) cells were collected and all E2-reactive (E2^+^) cells present were collected (316 to 11,613, median 1,461).

Determination of E2^+^ frequency was done using FlowJo software. Lymphocytes were downsampled to 500,000 so that cell numbers from all HCV subjects and healthy controls would be comparable. Some HCV subjects were sorted multiple times to increase yield for RNA sequencing and their E2^+^ frequencies were averaged.

### Isolation of RNA, library preparation, and sequencing

RNA was isolated using the RNeasy Micro Kit (Qiagen) according to the manufacturer’s instructions. Briefly, cells in lysis buffer containing 2-mercaptoethanol were combined with 70% ethanol and applied to a purification column. Following multiple washes and the application of DNAse, RNA was eluted in RNAse free water. RNA quality was verified using a 2100 Bioanalyzer (Agilent). cDNA libraries were produced using the SMARTer Human BCR IgG IgM H/K/L Profiling Kit (Takara) according to the manufacturer’s instructions and with the addition of unique molecular identifiers (UMIs). AMPure XP beads (Beckman) were used for purification steps. All sequencing was performed on an Illumina MiSeq at a depth of 1 million reads. cDNA library quality was verified using a 2100 Bioanalyzer (Agilent) and quantified using a Qubit (ThermoFisher). Clonotypes were quantified using MiXCR software^24^. Clonotypes were defined using sequences with nucleotides with Phred quality scores > 20, however subsequent sequences with nucleotides with lower quality scores could be matched to existing clonotypes if the number of low quality nucleotides was < 0.7%. All additional analysis was done in R Studio (version 4.0.2).

### Antibody synthesis and ELISA

Heavy chain variable sequences were synthesized (Twist Biosciences) and cloned into a human IgG1 expression plasmid (pTwist CMV BetaGlobin WPRE Neo_IgG1Fc). Kappa and lambda variable sequences were synthesized and cloned into a human IGK or IGL expression plasmid (pTwist CMV BetaGlobin WPRE Neo_Kappa_TAG or pTwist CMV BetaGlobin WPRE Neo_Lambda_TAG). Expi293 cells were transfected with equivalent concentrations of the heavy and light chain vectors using the Gibco Expi293 Expression System, as described by the vendor (Thermo Fisher Scientific). Ig was harvested and purified using Pierce™ Protein G Agarose (Cat. #20398) and buffer-exchanged into PBS, then concentrated using Pierce™ Protein Concentrator PES, 30K MWCO (Cat. #88529).

For E2 binding assays, immulon 2b microtiter plates were coated with Lectin followed by incubation with sE2 antigen (50 μL at 1 μg/mL) overnight. The next day, the plates were blocked with PBS-TMG (PBS + 0.5% tween 20+ 1% non-fat dry milk+1% goat serum) and then incubated with serial dilutions of our experimental mAb, a positive control mAb (HEPC74), or a negative control antibody (human IgG). Anti-human IgG-HRP (BD-Pharmingen Cat #555788) was used at 1:4000 dilution with TMB peroxidase substrate and 1N sulfuric acid to stop the reaction. Plates were read at an absorbance of 450nm.

### Statistical analysis

All statistical tests were performed in R Studio. Two group comparisons were performed with t tests if data were normally distributed (based on the Shapiro Wilk normality test) or Mann Whitney rank test if data were not normally distributed. Multi-group comparisons were performed using one-way ANOVA if data were normally distributed or Kruskal-Wallis test if data were not normally distributed, with p values adjusted for multiple comparisons using the Bonferroni method. Comparisons of proportions were done using Fisher’s exact test with p values adjusted for multiple comparisons using the Bonferroni method.

### Study approval

This research was approved by the Johns Hopkins University School of Medicine’s Institutional Review Board (IRB). Prior to blood collection, all participants provided informed written consent.

## Supporting information

Manuscript

## Data Availability

All data referred to in the manuscript is available on request.

## Acknowledgements

We thank all the participants in the BBAASH cohort and members of the Johns Hopkins Viral Hepatitis Center for thoughtful discussion. We thank the Bloomberg Flow Cytometry and Immunology Core for equipment and technical assistance, and Tricia Nilles, MS, and Hao Zhang. M.D. for the acquisition of sorted cells. We would like to thank the Sidney Kimmel Comprehensive Cancer Center Experimental and Computational Genomics Core for support with the BCR sequencing and analysis. This work was supported by National Institutes of Health grants R01AI127469 and R21AI151353 (to JRB) and U19AI159822 (to ALC and JRB).

