## Supplementary material for "Convergent antibody responses associated with broad neutralization of hepatitis C virus and clearance of infection": Manuscript

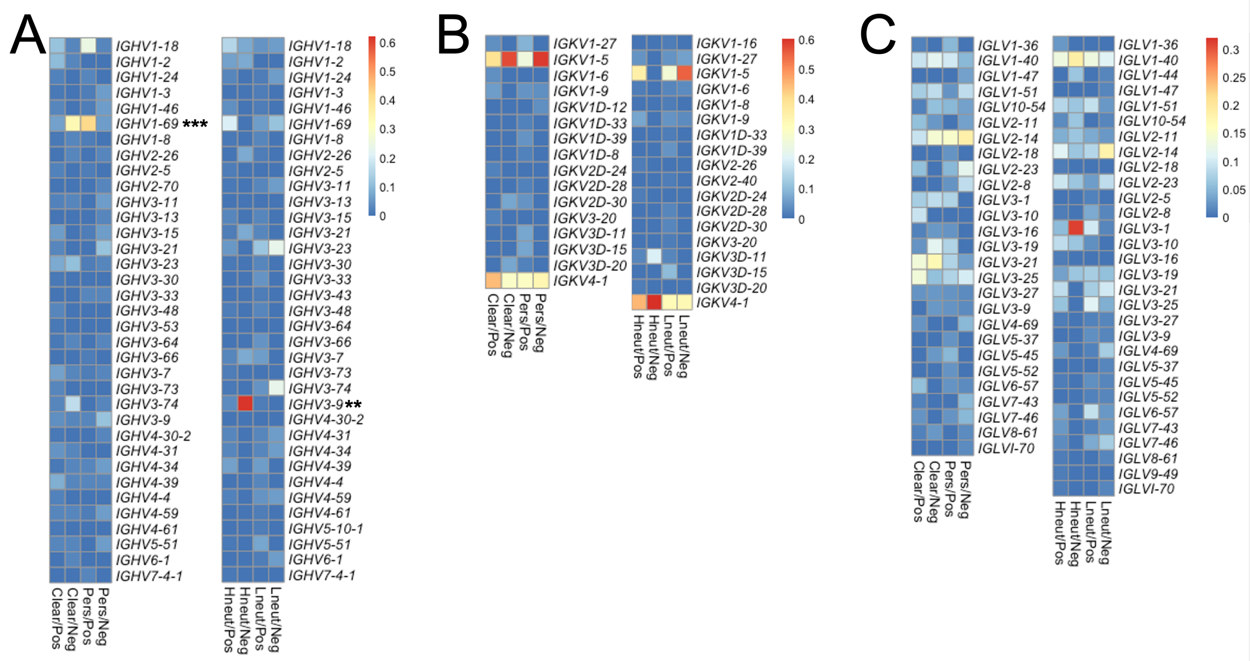


**Supplemental Figure 1. V-gene usage for group-specific public clonotypes.** Heatmap showing *IGHV* **(A)**, *IGKV* **(B)**, and *IGLV* **(C)** gene usage of the group-specific public clonotypes of the clearance/persistence (left) or high/low neutralization (right) groups. Usage of a given V-gene is expressed as a proportion of total V-gene usage by the group. Starred V-genes represent statistically significant differences between clearance/E2$^+$ and persistence/E2$^+$ or high neutralization/E2$^+$ and low neutralization/E2$^+$. All other comparisons were not significant. Statistical comparisons were made using Fisher's exact test with the Bonferroni correction for multiple comparisons. **, $P < 0.01$; ***, $P < 0.001$.

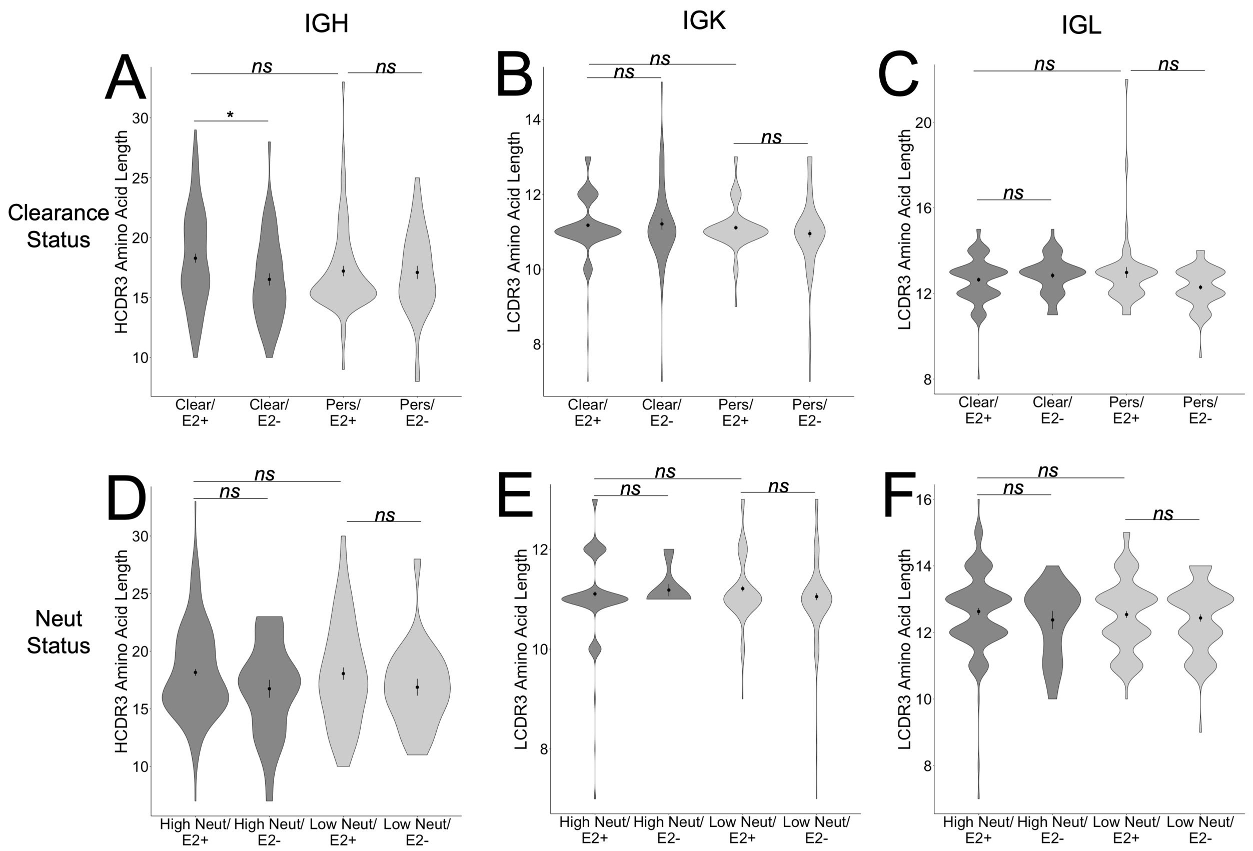


**Supplemental Figure 2. CDR3 length for group-specific public clonotypes.** Comparison of CDR3 lengths between the IGH **(A)**, IGK **(B)**, and IGL **(C)** group-specific public clonotypes of clearance and persistence groups. Comparisons are also shown of CDR3 lengths between the IGH **(D)**, IGK **(E)**, and IGL **(F)** group-specific public clonotype of high and low neutralization groups. Central dots and vertical lines represent means and standard errors, respectively. Statistical comparisons were made using Kruskal-Wallis test with the Bonferroni correction for multiple comparisons. *, P < 0.05; ns, not significant.
